# Determining Line of Therapy from Real-World Data in Non-Small Cell Lung Cancer

**DOI:** 10.1101/2024.02.22.24302979

**Authors:** Connor B. Grady, Wei-Ting Hwang, Joshua E. Reuss, Wade Iams, Amanda Cass, Geoffrey Liu, Devalben Patel, Stephen V. Liu, Gabriela Liliana Bravo Montenegro, Tejas Patil, Jorge J. Nieva, Amanda Herrmann, Kristen A. Marrone, Vincent K. Lam, William Schwartzman, Jonathan Dowell, Liza C Villaruz, Kelsey Leigh Miller, Jared Weiss, Fangdi Sun, Vamsidhar Velcheti, D. Ross Camidge, Charu Aggarwal, Lova Sun, Melina E. Marmarelis

**Affiliations:** Department of Biostatistics, Epidemiology and Informatics, Perelman School of Medicine, 423 Guardian Drive, Philadelphia, PA 19104; Department of Medicine Georgetown University, 3800 Reservoir Road NW, Washington, DC 20007; Division of Hematology/Oncology, Department of Medicine, Vanderbilt University Medical Center, 2220 Pierce Ave, 780 PRB, Nashville, TN 37232; Princess Margaret Cancer Centre, 610 University Ave, Toronto, ON M5G 2C4, Canada; University of Colorado Cancer Center, 1665 Aurora Court, MS F704, Aurora, CO 80045; University of Southern California/Norris Comprehensive Cancer Center, 1441 Eastlake Avenue NOR 3400, Los Angeles, CA 90033; Johns Hopkins University School of Medicine, 300 Mason Lord Drive, Baltimore, MD 21224; Harold C Simmons Comprehensive Cancer Center, UT Southwestern, 6202 Harry Hines, Blvd, 9 Floor, Dallas Texas, 75235; UPMC Hillman Cancer Center, 5150 Centre Avenue, 5 Floor, Pittsburgh, PA 15232; Lineberger Comprehensive Cancer Center, 170 Manning Drive, Room 3115, Chapel Hill, NC 27599; UCSF School of Medicine, University of California San Francisco, 533 Parnassus Ave, San Francisco, CA 94143; NYU Grossman School of Medicine, 550 1 Ave, New York, NY 10016; Department of Medicine, Division of Hematology and Oncology, Perelman School of Medicine, University of Pennsylvania, 3400 Civic Center Blvd, 10^th^ Floor, South Tower, Philadelphia, PA 19104

**Keywords:** Non-small cell lung cancer, EGFR, ALK, ROS1, real-world data

## Abstract

**Introduction:** Determining lines of therapy (LOT) using real-world data is crucial to inform clinical decisions and support clinical research. Existing rules for determining LOT in patients with metastatic non-small cell lung cancer (mNSCLC) do not incorporate the growing number of targeted therapies used in treatment today. Therefore, we propose rules for determining LOT from real-world data of patients with mNSCLC treated with targeted therapies.

**Methods:** LOT rules were developed through expert consensus using a real-world cohort of 550 patients with *ALK*+ or *ROS1*+ mNSCLC in the multi-institutional, electronic medical record-based Academic Thoracic Oncology Medical Investigators Consortium’s (ATOMIC) Driver Mutation Registry. Rules were subsequently modified based on a review of appropriate LOT determination. These resulting rules were then applied to an independent cohort of patients with *EGFR*+ mNSCLC to illustrate their use.

**Results:** Six rules for determining LOTs were developed. Among 1133 patients with *EGFR* mutations and mNSCLC, a total of 3168 regimens were recorded with a median of 2 regimens per patient (IQR, 1-4; range, 1-13). After applying our rules, there were 2834 total LOTs with a median of 2 LOTs per patient (IQR, 1-3; range, 1-11). Rules 1-3 kept 11% of regimen changes from advancing the LOT. When compared to previously published rules, LOT assignments differed 5.7% of the time, mostly in LOTs with targeted therapy.

**Conclusion:** These rules provide an updated framework to evaluate current treatment patterns, accounting for the increased use of targeted therapies in patients with mNSCLC and promote standardization of methods for determining LOT from real-world data.

**Key Points:**

- Use of targeted therapy to treat patients with mNSCLC is growing
- Determining lines of therapy from real-world data is crucial for clinical research
- Our rules aim to advance the line of therapy to reflect changes in clinical status
- Using these rules can lead to better method harmonization in mNSCLC research

## Introduction

When using real-world data to answer clinical questions in oncology, accurate determination of lines of therapy (LOT) using treatment records is crucial to conducting meaningful analyses. In the absence of detailed information on progression and toxicity, investigators rely on treatment records alone to infer changes in clinical disease status and determine LOTs. This non-uniform methodology may cause significant variation in how LOTs are defined depending on malignancy type, data source, and treatment patterns. Standardized rules and algorithms for determining LOTs are needed to improve the reproducibility and comparability of real-world data analyses.

There is tremendous opportunity for impactful analysis of real-world data from patients with metastatic non-small cell lung cancer (mNSCLC), but standardized LOT determination has been difficult. Systemic therapies for mNSCLC now include immunotherapy, chemotherapy, and targeted therapies, which can be used alone or in combination. Because of this, treatment patterns and sequencing are complex and heterogeneous^1^. This is particularly true for the subset of patients with mNSCLC and rare molecular characteristics and patients with co-morbidities where complex treatment patterns and outcomes are only captured outside of clinical trials.

Previously, others have proposed algorithms for determining LOTs for patients with solid tumors, lung cancer, and mNSCLC.^2^ However, these algorithms do not specifically incorporate common treatment patterns for patients with targetable alterations, such as *EGFR, ALK*, and *ROS1* alterations. In this report, we aim to propose new rules for determining LOT from real-world data of patients with mNSCLC, focusing on treatment patterns among patients receiving targeted therapies.

## Methods

We defined a regimen as any systemic anticancer agent (i.e., chemotherapy, immunotherapy, or targeted therapy) or combination of systemic agents administered at the same time. First, we developed rules for determining LOT (“LOT rules”) by expert consensus and review of existing literature. Next, these rules were applied to treatment records from 550 patients with mNSCLC and sensitizing *ALK* or *ROS1* fusions in the ATOMIC Driver Mutation Registry. ATOMIC is a multi-institutional collaborative of twelve academic centers across North America. Trained data abstractors used a standardized digital form to record patients’ regimen agent(s) and start/end dates. Finally, thoracic oncologist members of ATOMIC reviewed the LOT determination and revised the rules to reflect practice patterns most indicative of changes to clinical disease status. These modified rules were then applied to a separate ATOMIC cohort of 1,133 patients with mNSCLC and *EGFR* sensitizing mutations. Rules were also developed to identify LOTs that are most likely applicable to early-stage disease (“ESD rules”) since data sources of patients with mNSCLC may inadvertently include treatments for this disease stage.

In this report, we demonstrate the use of LOT and ESD rules on treatment records from the ATOMIC *EGFR* cohort. We compared our LOT determination with those of an existing algorithm for mNSCLC proposed by Hess et al using the SAS macro %mnsclc_lot^2^. Analyses were performed in R version 4.2.1^3^ and SAS software version 9.4. This study was approved by the University of Pennsylvania Institutional Review Board (IRB #829009).

## Results

We propose six rules for determining LOT in real-world data of patients with mNSCLC (**Table 1, Figure 1**). Among 1,133 patients in ATOMIC with *EGFR* alterations and mNSCLC, a total of 3168 regimens were recorded with a median of 2 regimens per patient (IQR, 1-4; range, 1-13). After applying our rules, there were 2834 total LOTs with a median of 2 LOTs per patient (IQR, 1-3; range, 1-11). Rules 1-3 prevented 334 regimen changes from advancing the LOT (**Table 1**). Thirty-seven patients switched regimens within 28 days of the start of the first LOT and had a line 0.5 recorded.

**Table 1:**
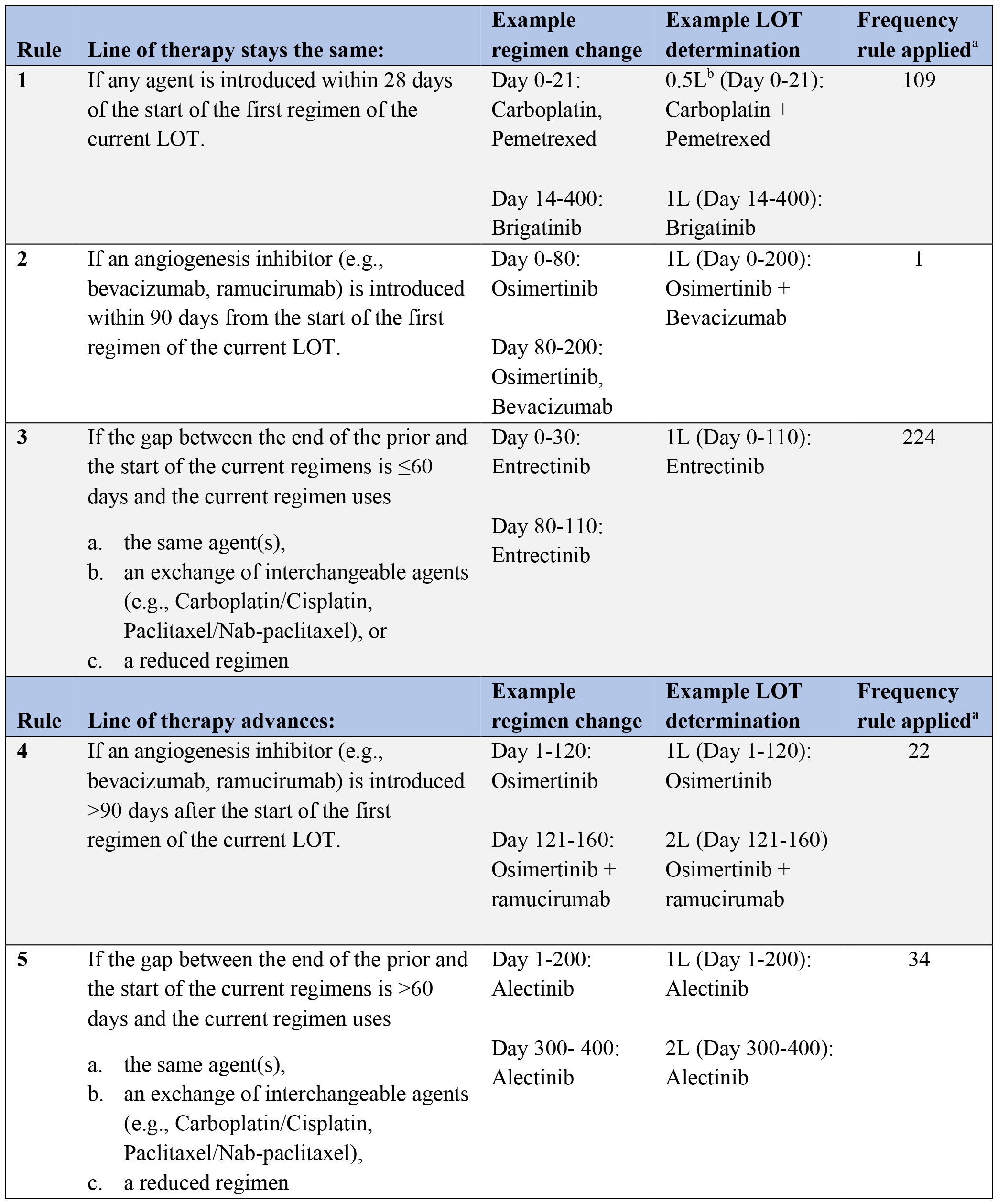
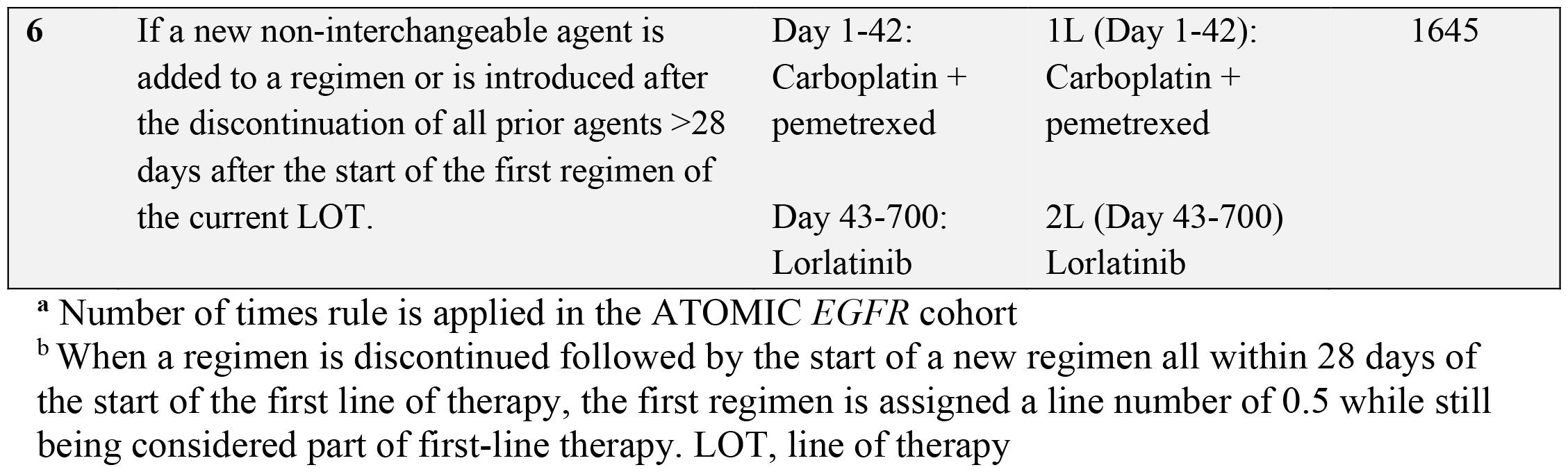
Proposed rules for line of therapy determination from real-world treatment regimens.

**Figure 1:**
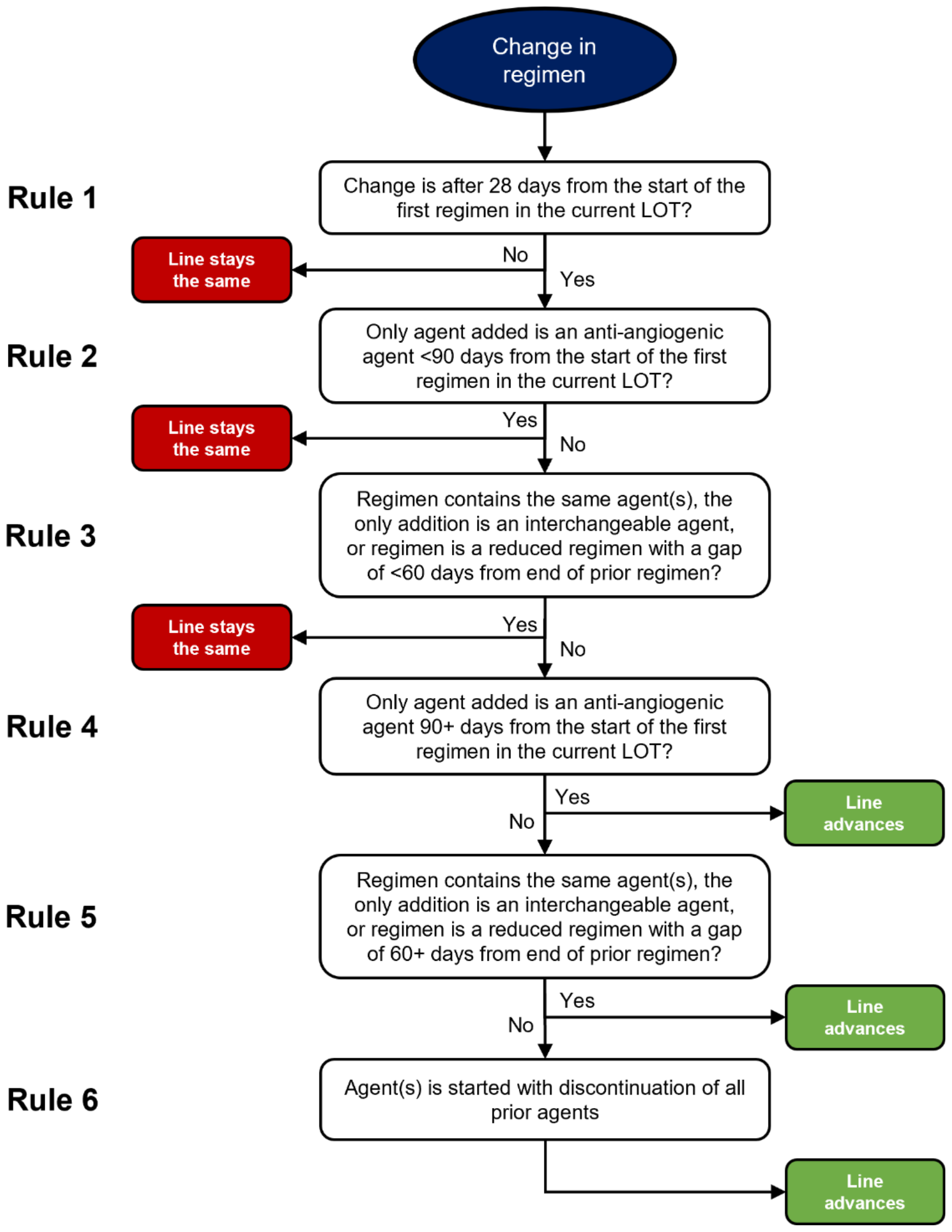
Algorithm for line of therapy determination.

Comparatively, after applying an algorithm proposed by Hess et al. to the same treatment records, there were 2771 total LOTs with a median of 2 per patient (IQR 1-3; range 1-9). Among our method’s 3999 agent-LOT assignments (e.g., osimertinib as 1L, carboplatin as 2L, etc.), ours differ from the Hess method in 5.7% of assignments (Table 2).

**Table 2:** Example line of therapy determination differences between the proposed rules and other (Hess et al.) rules.

| Patient | Duration of regimen (days) | Regimen agent(s) | Proposed LOT | Hess et al. LOT |
| --- | --- | --- | --- | --- |
| Patient A | 0-150 | Erlotinib | 1 | 1 |
|  | 151- 242 | Osimertinib | 2 | 2 |
|  | 243-438 | Bevacizumab, Carboplatin, Pemetrexed | 3 | 3 |
|  | 439-674 | Pemetrexed | 3 | 3 |
|  | 675-702 | Bevacizumab, Pemetrexed | 4 | 3 |
|  | 703-849 | Pemetrexed | 4 | 3 |
|  | 881-ongoing | Bevacizumab, Erlotinib | 5 | 3 |
| Patient B | 0-630 | Bevacizumab, Erlotinib | 1 | 1 |
|  | 632-1036 | Osimertinib | 2 | 2 |
|  | 1038-1091 | Osimertinib, Crizotinib | 3 | 2 |
| Patient C | 0 | Bevacizumab, Carboplatin, Pemetrexed | 1 | 1 |
|  | 30-800 | Erlotinib | 2 | 2 |
|  | 802-904 | Bevacizumab, Carboplatin, Pemetrexed | 3 | 3 |
|  | 905-1010 | Bevacizumab, Pemetrexed | 3 | 3 |
|  | 1205-1849 | Osimertinib | 4 | 4 |
|  | 1851-2035 | Osimertinib, Gefitinib | 5 | 4 |

Highlighted cells indicate differences in how lines of therapy are determined between the proposed rules and other (Hess et al.) rules. LOT, line of therapy.

Additionally, we developed two rules to remove systemic therapy agents likely used in the treatment of early-stage disease (ESD) from the LOT determination. (1) If a LOT was durvalumab monotherapy administered prior to November 11, 2022 (i.e., prior to Food and Drug Administration approval for durvalumab in stage IV NSCLC)^4^ or (2) if the LOT was chemotherapy with a cumulative cycle duration of 30-140 days and no subsequent LOT for at least four months of follow-up, the LOT should be considered as a treatment for early-stage disease and excluded from the patient’s LOT enumeration for mNSCLC. Among patients in ATOMIC with *EGFR* alterations and mNSCLC, the first LOT for 81 patients met the criteria for early-stage disease treatment (ESD Rule 1 met for two patients and ESD Rule 2 met for 79). These LOTs were removed, and the remaining LOTs were renumbered as LOTs for mNSCLC.

## Discussion

We propose six simple rules to determine whether a change in treatment for mNSCLC should or should not advance the LOT in real-world data. Our LOT assignments led to different classifications from previous work in 5.7% of cases, primarily in characterizing targeted therapy-containing regimens. Since LOT assignment can affect outcome measures such as real-world progression-free survival or time to treatment discontinuation, even small differences may lead to different conclusions in an observational study. Therefore, it is critical that the LOT determination accurately reflects clinical practice.

There are several ways that our proposed rules better characterize LOTs in mNSCLC populations treated with targeted therapy compared to previously reported methods (**Supplemental table 1**).^2,4–7^ First, we distinguish between biologics and therapies targeting oncogenic drivers in mNSCLC. This distinction is important because biologics, such as angiogenesis inhibitors, may have little anti-tumor activity alone and are often added or removed from treatment regimens without a clear change in disease status, while targeted therapies have tremendous anti-tumor activity and are used in the setting of a change in disease status. Second, there are instances where targeted therapies are added to or replace a previous regimen because of a newly identified targetable mutation and not because of a change in disease status. The rules proposed allow for this approach by allowing for 1 cycle of chemotherapy to be given while waiting for molecular testing results prior to switching to a targeted therapy without advancing the LOT. Third, we allow for a treatment pause and re-initiation within 60 days to be considered the same LOT, which may be more appropriate for targeted therapies typically taken daily by mouth that are paused and restarted for toxicity.^5–7^

Retrospective determination of whether a treatment was given in the setting of metastatic disease versus early-stage disease is critical to determining real-world clinical endpoints. In this historical dataset, we used rules based on the approved therapies at the time of the dataset, but even while completing the analysis for this study the indications for several systemic therapies expanded to include early-stage disease, making the agent itself an unreliable indicator of early-stage treatment. Given this complexity, extracting the date of metastatic diagnosis at the time of data abstraction will become even more important to evaluate the practice patterns contemporary to the time of the dataset.

We believe wider use of this framework for LOT determination could lead to better method harmonization of observational studies in mNSCLC and allow for comparison and aggregation across datasets in the future.

## Data Availability

Data produced in the present study are available upon reasonable request to the authors.

## Acknowledgements

This work is supported in part by grants from Takeda, AstraZeneca and LUNGevity.

## Tables and figures

**Supplemental table 1:** Differences between proposed rules and Hess et al. rules.

| Scenario | Line of therapy advances |  |
| --- | --- | --- |
|  | Proposed rules | Hess et al. rules |
| Targeted or immunotherapy is added to a regimen 28+ days after the start of the first regimen in the current LOT. | Yes | No |
| Targeted or immunotherapy is started with discontinuation of all agents in prior regimen. | Yes*<br>*Except if started <28 days from start of LOT | Yes |
| An interchangeable agent is exchanged 60+ days after the end of the prior regimen. | Yes | No |
| Introduction of an anti-angiogenic agent <90 days from the start of any LOT, with or without discontinuation of the prior regimen. | No | Yes*<br>*Except if pemetrexed is part of prior regimen and given with anti-angiogenic agent in the first LOT |
| A regimen is reduced but with a gap in treatment of 60+ days.<br>(e.g., Day 0-60: Carboplatin + Alectinib, Day 140-ongoing: Alectinib) | Yes | Not addressed in rules |

LOT, line of therapy

